# A Genotype-Guided P2Y12-Inhibitor De-Escalation Strategy in Acute Coronary Syndrome: Observational Evidence from the POPULAR-GUIDE PCI

**DOI:** 10.1101/2025.08.19.25334023

**Authors:** W.W.A. van den Broek, Jaouad Azzahhafi, Qiu Ying Y.F van de Pol, Dean R.P.P. Chan Pin Yin, Niels M.R. van der Sangen, Shabiga Sivanesan, J. Peper, Ankie M. Harmsze, Ronald J. Walhout, Melvyn Tjon Joe Gin, Nicoline J. Breet, Jorina Langerveld, Yolande Appelman, Ron H.N. van Schaik, José P.S. Henriques, Wouter J. Kikkert, Jurriën M. ten Berg

## Abstract

**Background and Aims:** A genotype-guided de-escalation strategy - switching from a potent P2Y12-inhibitor to clopidogrel - may represent an effective and safe approach to reducing bleeding risk in patients with acute coronary syndrome (ACS). This analysis aimed to evaluate the safety and effectiveness of routine genetic testing to guide antiplatelet therapy in clinical practice.

**Methods:** In this investigator-initiated, prospective, multicentre implementation study, patients were divided into a standard care cohort, where antiplatelet therapy was prescribed at the physician’s discretion (with a potent P2Y12 inhibitor as the default choice), and a genotype-guided cohort. In the genotype-guided group, physicians were recommended to switch to clopidogrel in noncarriers of *CYP2C19* loss-of-function alleles during hospital admission. The primary endpoints were major adverse cardiac events (MACE), defined as a composite of cardiovascular death, myocardial infarction, or stroke, and major or non-major clinically relevant bleeding (Bleeding Academic Research Consortium types 2, 3, or 5), at one year of follow-up. Hazard ratios were adjusted for baseline differences between cohorts using multivariable Cox regression.

**Results:** A total of 9,907 patients were included in the analysis. Of these, 1,208 (12%) were included in the genotype-guided cohort, while 8,699 (88%) were assigned to the standard care cohort. MACE occurred in 107 patients (8.9%) in the genotype-guided cohort and 897 patients (10.3%) in the standard care cohort (_adj_HR 1.05; 95% CI 0.85-1.29; *P* = 0.64). Major or non-major clinically relevant bleeding was reported in 146 patients (12.1%) in the genotype-guided cohort compared to 1,384 patients (15.9%) in the standard care cohort (_adj_HR 0.79; 95% CI 0.67–0.94; *P* = 0.01).

**Conclusion:** In patients with ACS receiving antiplatelet therapy, implementation of a CYP2C19 genotype-guided de-escalation strategy in clinical practice significantly reduced major and non-major clinically relevant bleeding compared to standard care at 12 months, without increasing ischemic events.

## Introduction

Acute coronary syndrome (ACS) continues to be a significant cause of morbidity and mortality worldwide, necessitating effective strategies to reduce ischemic events in affected patients.[1] Dual antiplatelet therapy (DAPT), consisting of aspirin and a P2Y12-inhibitor, is the cornerstone of secondary prevention in these patients.[2] While the more potent P2Y12-inhibitors ticagrelor and prasugrel offer enhanced protection against ischemic events, their use is associated with an increased risk of bleeding, a complication that significantly impacts patient outcomes and is associated with increased morbidity and mortality.[3,4]

The challenge of balancing ischemic and bleeding risks has driven the development of personalized treatment strategies, including the adoption of de-escalation strategies.[5] These strategies involve switching from potent P2Y12-inhibitors to less potent alternatives, such as clopidogrel, to mitigate bleeding risks without compromising ischemic protection. A promising approach to optimize this balance in patients with ACS is a genotype-guided de-escalation strategy. Genetic testing for the *CYP2C19* genotype, which encodes the enzyme essential for clopidogrel activation, enables tailored treatment by identifying patients with loss-of-function (LOF) alleles. These individuals are more likely to experience high on-treatment platelet reactivity (HTPR) to clopidogrel, which in turn is associated with increased ischemic risk.[6] Patients without these genetic variants (normal metabolizers), who comprise approximately 70% of the European population, may safely switch to clopidogrel, reducing both bleeding risks and healthcare costs.[7,8] The POPular Genetics trial demonstrated the efficacy and safety of a *CYP2C19* genotype-guided de-escalation strategy in a randomized setting, showing reduced bleeding without an evident increase in ischemic events among patients with ST-segment elevation myocardial infarction (STEMI).[9] Despite these findings and recommendations from various expert opinion groups, the implementation of this strategy in clinical practice remains low, resulting in limited evidence from real-world settings.[10–12] The controlled nature of clinical trials often excludes high-risk populations and does not account for the complexities of routine clinical practice, raising questions about the generalizability of these results.

To address this gap, our study examines the implementation of a genotype-guided de-escalation strategy in a real-world ACS population. This analysis aims to provide insights into the safety and effectiveness of routine genetic testing to guide antiplatelet therapy, focusing on bleeding and ischemic outcomes in an all-comers cohort at one year follow-up. By bridging the evidence gap between clinical trials and real-world practice, our findings aim to inform broader adoption of personalized antiplatelet therapy strategies.

## Methods

### Study design

The POPular GUIDE PCI was an investigator-initiated, prospective, observational, multicentre study. It was conducted as an implementation initiative within the ongoing FORCE-ACS (Future Optimal Research and Care Evaluation in Patients with Acute Coronary Syndrome) Registry (NCT03823547), which includes nine non-interventional and interventional cardiac centres in the Netherlands.[13] The research protocol of the FORCE-ACS registry was approved by the institutional review boards of all participating medical centres. Enrolment procedures have been prescribed previously.[13] All patients included in the registry have provided either written or digital informed consent.

### Study Population and Data Collection

Patients aged 18 years or older are eligible for enrolment in the FORCE-ACS registry upon admission with a suspected ACS, including unstable angina, non-ST-elevation myocardial infarction (NSTEMI), or STEMI. The registry has no exclusion criteria. Data was prospectively collected by members of the investigation team during a follow-up of three years. Follow-up was conducted via electronic health record (EHR) review and questionnaires administered at predefined intervals: 1, 12, 24, and 36 months after admission. The questionnaires included specific questions about ischemic and bleeding events, assessed quality of life, and gathered information on drug use and any changes in antiplatelet therapy.

### Treatment procedures

Since 2015, patients with (suspected) ACS were consecutively enrolled in the FORCE-ACS registry. Antithrombotic treatment followed local protocols, comprising a loading dose of aspirin, a potent P2Y12-inhibitor, and heparin, in line European Society of Cardiology guidelines.[14] In June 2021, the implementation of the genotype-guided de-escalation strategy was initiated using on-site testing facilities in the St. Antonius hospital, Nieuwegein, the Netherlands. Details on the genotype guided de-escalation have been described previously.[15] Initially, CYP2C19 genotyping was performed using the Genomadix point-of-case assay which required buccal swap samples. In March 2022, the implementation protocol was updated to include on-site *CYP2C19* genotyping using genomic DNA extracted from venous blood in the central laboratory. The local protocol required that every patient with ACS underwent *CYP2C19* genotype testing immediately after admission, with STEMI patients primarily tested via the point-of-care system and NSTEMI patients predominantly tested in the central laboratory. Patients carrying at least one *CYP2C19* LOF allele (*2 or *3), categorized as intermediate (IM) or poor metabolizers (PM), remained on ticagrelor or prasugrel. In non-carriers ([ultra-]rapid [UM/RM] or normal metabolizers [NM]), attending physicians received an automated notification to switch antiplatelet therapy from ticagrelor to clopidogrel, starting with a 600 mg loading dose followed by 75 mg daily. Ultimate prescribing decisions were left to the physician’s discretion.

As of 2023, three of the other participating sites adopted off-site genetic testing using blood samples, following the same de-escalation protocol. At these sites, de-escalation to clopidogrel was performed during the patient’s next follow-up visit.

### Study endpoints

The study had two primary endpoints: major adverse cardiac events (MACE), defined as a composite of cardiovascular death, myocardial infarction (MI), or stroke within 12 months of hospital admission, and major or non-major clinically relevant bleeding, classified according to the Bleeding Academic Research Consortium (BARC) types 2, 3, or 5. The secondary endpoint, net adverse cardiac events (NACE), was defined as a composite of all-cause death, MI, stroke, stent thrombosis and major bleeding (BARC 3 or 5). All the individual components of the primary endpoints were adjudicated by coordinating investigators of the FORCE-ACS registry.

P2Y12-inhibitor adherence was assessed by classifying medication changes into two groups: alterations, defined as any change in the prescribed P2Y12-inhibitor after hospital discharge, and disruptions, defined as discontinuation of P2Y12-inhibitor therapy for more than 14 days. Reasons for drug alterations were also documented.

### Statistical analysis

Patients were categorized into two cohorts: a standard care cohort, where P2Y12-inhibitors were prescribed at the treating physician’s discretion, and a genotype guided cohort, where patients underwent *CYP2C19* genotype testing with treatment recommendations tailored to the test results. Patients, in whom the *CYP2C19* genotype was already known at admission, were also included in the genotype-guided group. For this analysis, only patients with a final diagnosis of ACS, for whom antiplatelet therapy was indicated, were included.

Although this study is observational and based on ongoing registry data, a pre-specified sample size calculation was performed to ensure adequate power for detecting clinically meaningful differences between groups. We estimated the required sample size based on an assumed incidence of major or non-major clinically relevant bleeding of 11.0% in the standard-treatment group and 8.0% in the genotype-guided group, derived from a prior pilot analysis and the results of the POPular Genetics.[16] Using an alpha level of 0.05 and a power of 80%, we calculated that 1,125 patients in the genotype-guided group would be necessary to demonstrate superiority. To account for a 5% attrition rate, the final planned sample size was set at 1,181.

Continuous variables were summarized as medians with interquartile ranges (IQR) or means with standard deviations (SD), while categorical variables were presented as frequencies and percentages. Comparisons between the standard care and genotyped cohorts were conducted using Mann-Whitney U tests or t-tests for continuous variables and Chi-square or Fisher’s Exact tests for categorical variables. The primary analysis utilized a Cox proportional hazards model to estimate hazard ratios (HRs) with 95% confidence intervals. Potential confounders were included in the multivariable model based on clinical relevance (age, hypercholesterolemia, prior myocardial infarction, prior CABG, prior stroke, peripheral artery disease, atrial fibrillation, discharge diagnosis, use of diuretics, triple therapy, ACE inhibitors or ATII receptor blockers, proton pump inhibitors, PCI during admission, high bleeding risk, and complex PCI). Similar Cox proportional-hazards models were used for the analysis of subgroups defined according to age (<75 years or ≥75 years), sex (male or female), diabetes mellitus (yes/no), renal function (<60 or >60 mL/min/1.73 m²), clinical presentation (NSTEMI, or STEMI), high bleeding risk (Predicting Bleeding Complications in Patients Undergoing Stent Implantation and Subsequent Dual Antiplatelet Therapy [PRECISE-DAPT] <25 or ≥25) and complex PCI (yes/no), defined as the use of three or more stents, treatment of three or more lesions, stent length exceeding 60 mm, left main stenting, bifurcation stenting, or prior stent thrombosis. Time-to-event analyses were performed using Kaplan-Meier curves. The proportional hazards assumption was assessed using Schoenfeld residuals.

To account for baseline differences and reduce confounding in the comparison of outcomes, propensity score matching was performed using clinically relevant covariates. Matching followed a one-to-three protocol without replacement, applying the nearest neighbour method with a calliper of 0.2 standard deviations of the logit of the propensity score. Additional sensitivity analyses were conducted to refine the assessment of the genotype-guided strategy’s treatment effect in three different populations, one excluding patients treated with oral anticoagulants at discharge, one excluding those patients in whom the recommended choice of P2Y12-inhibitor based on the genetic test result was not followed and one only including patients that underwent PCI during admission.

Patients were evaluated from hospital admission until death, withdrawal of consent, or the last contact date. All statistical analyses were conducted R studio version 3.6.1 (Vienna, Austria).

## Results

### Patient characteristics

A total of 9,907 patients, enrolled between January 2015 and December 2023, were included in the analysis (**Figure 1**). Of these, 1,208 (12%) were included in the genotype-guided cohort, while 8,699 (88%) comprised the standard care cohort. The mean age of the overall population was 66 years (SD ±11.8), 2,827 (29%) were women, and 70% (N = 6,896) underwent PCI during hospital admission. Patients in the genotype-guided cohort exhibited a higher prevalence of prior bleeding events and STEMI as the index diagnosis, whereas the standard care cohort had a greater prevalence of prior MI, prior PCI, prior CABG, prior stroke, atrial fibrillation and peripheral artery disease (**Table 1**).

**Figure 1.**
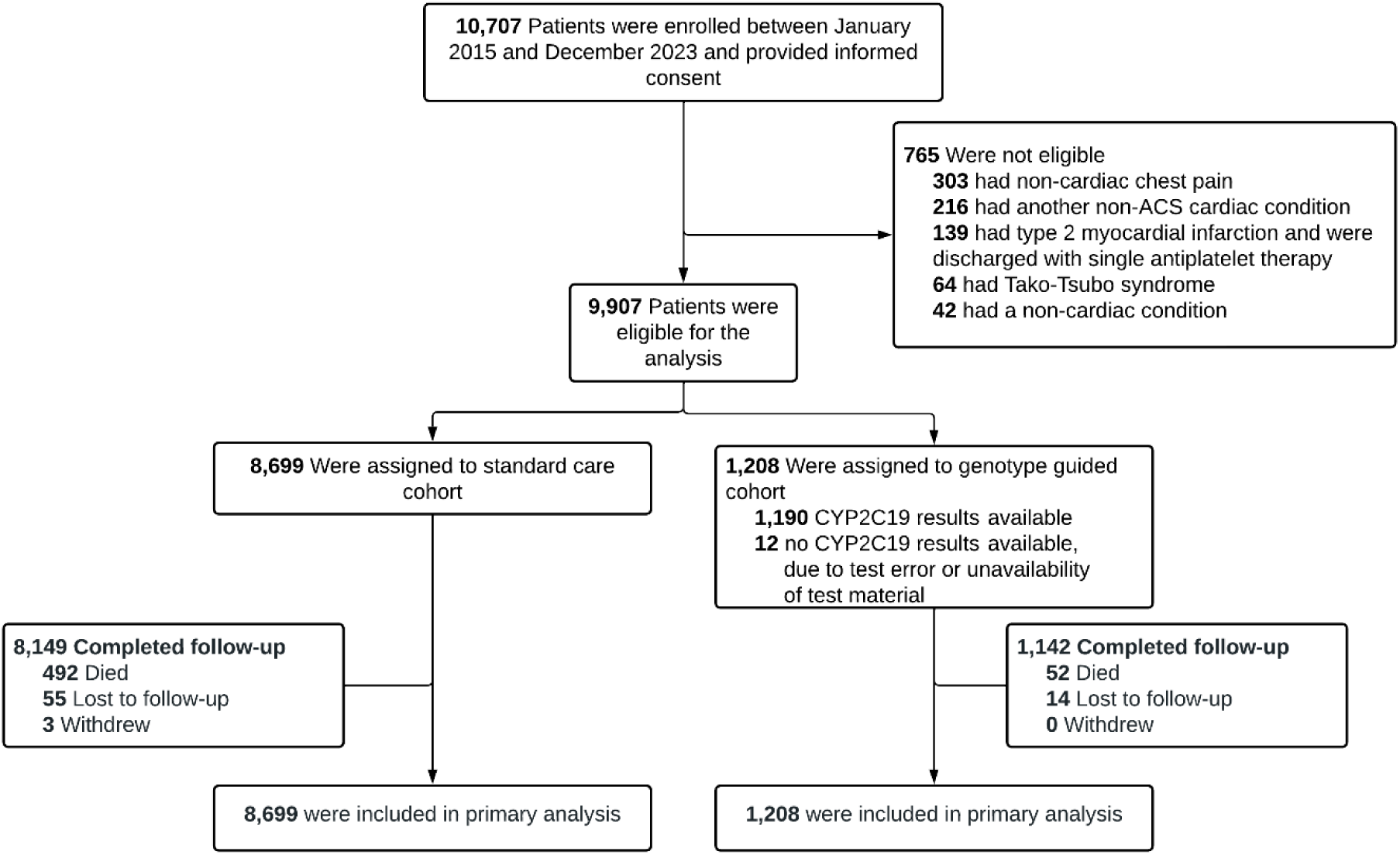
Study Flowchart. ACS = acute coronary syndrome

**Table 1.** Baseline table for the genotype guided cohort compared to the standard care cohort.

| Characteristics | Genotype Guided Cohort<br>(N = 1,208) | Standard Care cohort<br>(N = 8,699) | p-value |
| --- | --- | --- | --- |
| Age (mean [SD]) | 65.51 (11.39) | 66.43 (11.90) | 0.012 |
| Female sex (%) | 318 (26.3) | 2509 (28.8) | 0.075 |
| BMI (mean [SD]) | 27.68 (4.79) | 27.41 (4.44) | 0.057 |
| CYP2C19 LOF carrier (%) | 370 (30.6) | - | - |
| Hypertension (%) | 644 (53.6) | 4741 (55.9) | 0.139 |
| Dyslipidaemia (%) | 960 (79.5) | 4736 (54.4) | <0.001 |
| Diabetes Mellitus (%) | 245 (20.3) | 1801 (20.9) | 0.696 |
| Previous MI (%) | 197 (16.3) | 1788 (20.6) | 0.001 |
| Previous PCI (%) | 208 (17.2) | 1764 (20.3) | 0.014 |
| Previous CABG (%) | 67 (5.5) | 684 (7.9) | 0.005 |
| Prior stroke (%) | 82 (6.8) | 774 (8.9) | 0.017 |
| Atrial fibrillation (%) | 44 (3.6) | 713 (8.2) | <0.001 |
| PAD (%) | 55 (4.6) | 660 (7.6) | <0.001 |
| Kidney failure (%) | 41 (3.4) | 329 (3.8) | 0.558 |
| Previous bleeding event (%) | 127 (11.0) | 500 (5.9) | <0.001 |
| GRACE-score >140 (%) | 190 (15.7) | 1430 (16.4) | 0.559 |
| PRECISE-DAPT >25 (%) | 320 (26.6) | 2606 (30.4) | 0.008 |
| Discharge diagnosis |  |  | <0.001 |
| STEMI (%) | 628 (52.0) | 3443 (39.6) |  |
| NSTEMI (%) | 458 (37.9) | 4163 (47.9) |  |
| IAP (%) | 59 (4.9) | 702 (8.1) |  |
| **Treatment** |  |  |  |
| CAG during admission (%) | 1163 (96.4) | 8242 (94.8) | 0.021 |
| PCI during admission (%) | 905 (74.9) | 5991 (68.9) | <0.001 |
| DES stent (%) | 838 (69.4) | 5532 (63.6) | <0.001 |
| CAG access site |  |  | 0.007 |
| Radial (%) | 963 (79.7) | 6706 (77.1) |  |
| Femoral (%) | 189 (15.6) | 1402 (16.1) |  |
| Brachialis (%) | 1 (0.1) | 32 (0.4) |  |
| Vessel disease |  |  |  |
| 0-VD (%) | 53 (4.4) | 546 (6.3) | 0.012 |
| 1-VD (%) | 478 (39.6) | 2689 (30.9) | <0.001 |
| 2-VD (%) | 342 (28.3) | 1740 (20.0) | <0.001 |
| 3-VD (%) | 280 (23.2) | 1648 (18.9) | 0.001 |
| Graft dysfunction (%) | 22 (1.8) | 239 (2.7) | 0.074 |
| Bifurcation stenting (%) | 33 (3.3) | 119 (1.7) | <0.001 |
| Left main stenting | 29 (2.4) | 144 (1.7) | 0.064 |
| Complex PCI (%) | 172 (14.2) | 1010 (11.6) | 0.008 |
| Aspirin (%) | 992 (82.1) | 7420 (85.3) | 0.004 |
| Clopidogrel (%) | 723 (60.0) | 2505 (28.8) | <0.001 |
| Prasugrel (%) | 3 (0.3) | 46 (0.6) | 0.177 |
| Ticagrelor (%) | 422 (34.9) | 5494 (63.2) | <0.001 |
| Vitamin-K antagonist (%) | 46 (3.8) | 654 (7.5) | <0.001 |
| DOAC (%) | 134 (11.1) | 779 (9.0) | 0.019 |
| Oral anticoagulation (%) | 180 (14.9) | 1432 (16.5) | 0.182 |
| Triple therapy (%) | 8 (0.7) | 407 (4.7) | <0.001 |
| Beta-blocker (%) | 866 (71.7) | 6171 (70.9) | 0.59 |
| ACE inhibitor or AT-II antagonist (%) | 827 (68.5) | 6342 (72.9) | 0.001 |
| Lipid lowering drugs (%) | 1117 (92.5) | 7920 (91.0) | 0.102 |
| PPI (%) | 1127 (93.3) | 7314 (84.1) | <0.001 |

Data are n (%) unless stated otherwise. Missing data were observed for BMI (4.3%), access site (6.3%) and bifurcation stenting (19.3%); for all other variables, missing data were less than 3%. ACE = angiotensin-converting enzyme; AT-II = angiotensin-II; IQR = interquartile range; BMI= body mass index; CABG = coronary artery bypass grafting; CAG = coronary angiography; DES = drug eluting stent; CV = cardiovascular; IQR = interquartile range; kg = kilogram; MI = myocardial infarction; PAD = peripheral arterial disease; PCI = percutaneous coronary intervention; SD = standard deviation; VD = vessel disease.

### Treatment and Management

Coronary angiography was performed in 96% of genotype-guided patients and 95% of standard care patients (**Table 1**). PCI was more frequently performed in the genotype-guided cohort (75%) compared to the standard care cohort (69%). More patients in the genotype-guided cohort underwent a complex PCI-procedure (11.6% vs. 14.2%, *P* = 0.008).

Among the genotype-guided cohort, 69% of patients (838 of 1,208) were identified as non-carriers. At discharge, clopidogrel was prescribed to 60% of patients in the genotype-guided cohort (N = 723) and ticagrelor in 35% (N=422). In the standard care cohort, 29% of patients were treated with clopidogrel (N=2,505) and 63% with ticagrelor (N=5,494). Among loss-of-function carriers in the genotype-guided group (n = 370), 312 (84%) were treated with ticagrelor or prasugrel, while the remainder predominantly received clopidogrel, mainly due to concomitant oral anticoagulant use.

The rate of P2Y12 treatment alterations was significantly lower in the genotype-guided cohort compared to the standard care cohort (98 [8.1%] vs. 1243 [14.3%], *P* < 0.001). Similarly, dyspnoea leading to treatment alterations was reported less frequently in the genotype-guided cohort (36 [3.0%] vs. 517 [5.9%], P < 0.001). No significant difference was observed in treatment disruption between the two groups (36 [2.4%] vs. 517 [3.2%], *P* = 0.236).

### Comparison of Central Laboratory and Point-Of-Care Testing

The median time from hospitalization to test result was significantly shorter for patients tested using point-of-care testing (POCT) compared to the central laboratory (0 days [IQR 0–1] vs. 2 days [IQR 2– 4], *P* < 0.001, **Table 2**). Similarly, the median time from hospitalization to de-escalation was shorter with POCT (1 day [IQR 1–2] vs. 3 days [IQR 2–6], *P* < 0.001).

**Table 2.**
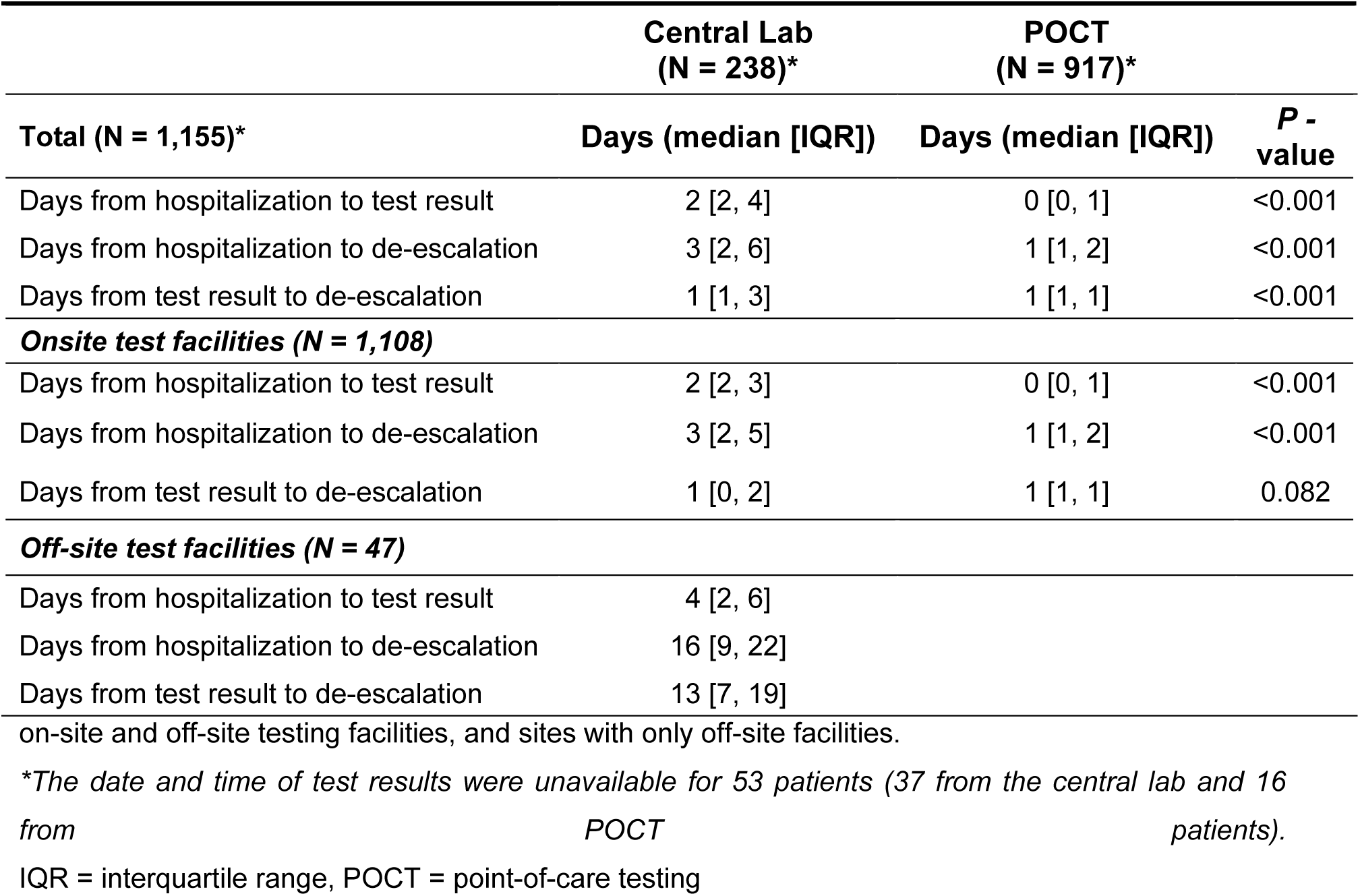
Comparison of central laboratory and point-of-care testing across between sites with both.

|  | Central Lab<br>(N = 238)* | POCT<br>(N = 917)* |  |
| --- | --- | --- | --- |
| Total (N = 1,155)* | Days (median [IQR]) | Days (median [IQR]) | P -<br>value |
| Days from hospitalization to test result | 2 [2, 4] | 0 [0, 1] | <0.001 |
| Days from hospitalization to de-escalation | 3 [2, 6] | 1 [1, 2] | <0.001 |
| Days from test result to de-escalation | 1 [1, 3] | 1 [1, 1] | <0.001 |
| **Onsite test facilities (N = 1,108)** |  |  |  |
| Days from hospitalization to test result | 2 [2, 3] | 0 [0, 1] | <0.001 |
| Days from hospitalization to de-escalation | 3 [2, 5] | 1 [1, 2] | <0.001 |
| Days from test result to de-escalation | 1 [0, 2] | 1 [1, 1] | 0.082 |
| **Off-site test facilities (N = 47)** |  |  |  |
| Days from hospitalization to test result | 4 [2, 6] |  |  |
| Days from hospitalization to de-escalation | 16 [9, 22] |  |  |
| Days from test result to de-escalation | 13 [7, 19] |  |  |

on-site and off-site testing facilities, and sites with only off-site facilities.
\*The date and time of test results were unavailable for 53 patients (37 from the central lab and 16 from POCT patients).
IQR = interquartile range, POCT = point-of-care testing

At the site with on-site testing facilities, POCT demonstrated faster turnaround times, with a median of 0 days (IQR 0–1) from hospitalization to test result compared to 2 days (IQR 2–3) for the central laboratory (*P* < 0.001). De-escalation also occurred more rapidly for POCT patients (median 1 day [IQR 1–2]) compared to those tested via the central laboratory (3 days [IQR 2–5], *P* < 0.001). At the sites with off-site testing facilities only, delays were more pronounced with central laboratory testing. The median time from hospitalization to test result was 4 days (IQR 2–6), and de-escalation occurred at a median of 16 days (IQR 9–22).

### Clinical Outcomes

#### Primary outcomes

During the median follow-up duration of 365 days (mean 348 days, IQR: 365–365 days), MACE occurred in 107 of 1,208 patients (8.9%) in the genotype guided cohort and 897 of 8,699 patients (10.3%) in the standard care cohort (_adj_HR, 1.05; 95% confidence interval [CI], 0.85-1.29; *P* = 0.64) (**Figure 2A** and **Table 3**). Major or non-major clinically relevant bleeding occurred in 146 patients (12.1%) in the genotype guided cohort and 1,384 patients (15.9%) in the standard care cohort (_adj_HR, 0.79; 95% CI, 0.67-0.94; P = 0.01, **Figure 2B**), with consistent results observed across other bleeding classifications (**Supplementary Appendix Table 1**). Kaplan-Meier analysis demonstrated similar results, showing no significant difference in MACE, but a highly significant reduction in major or non-major clinically relevant bleeding (log-rank *P* = 0.0006).

**Figure 2.**
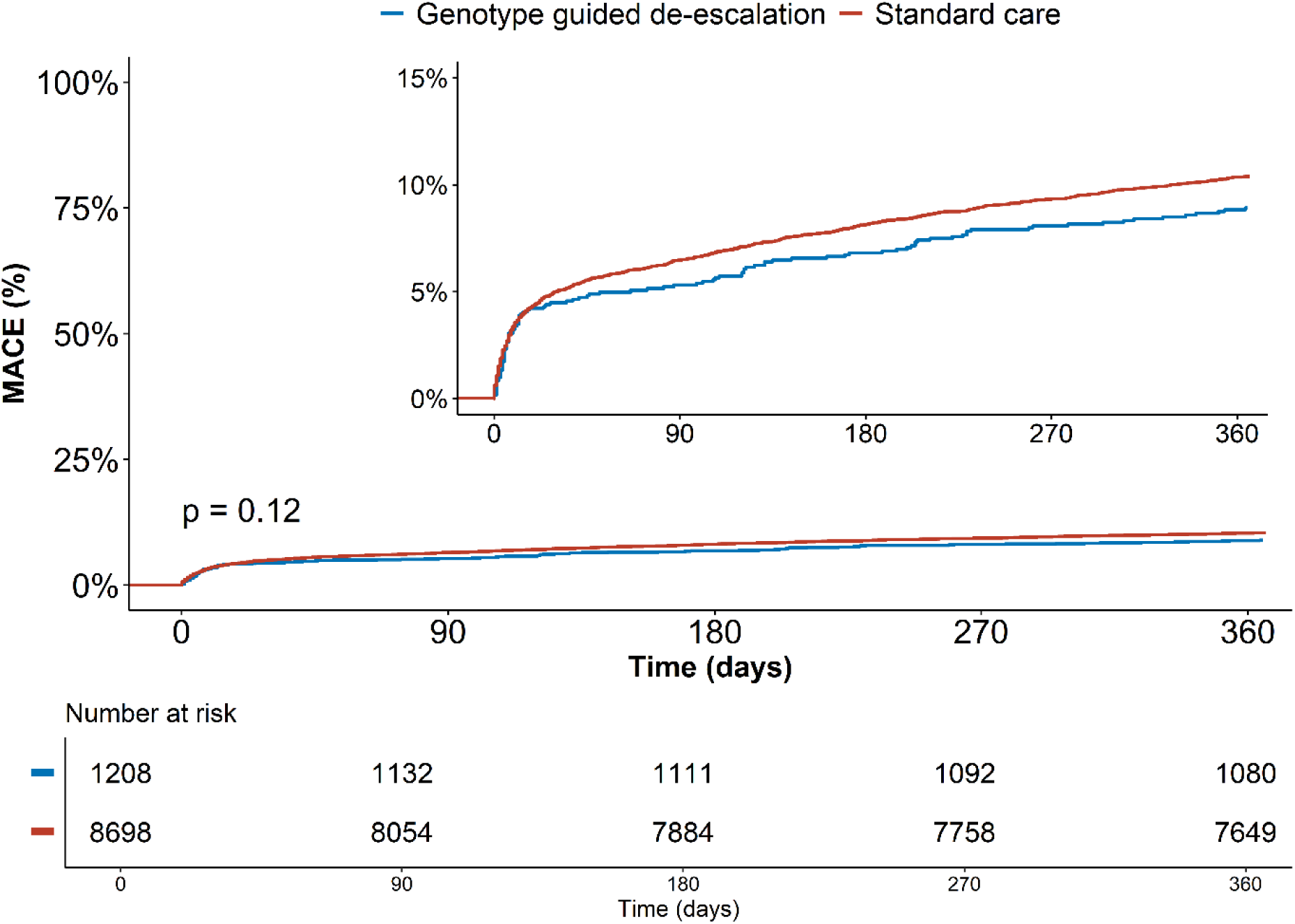
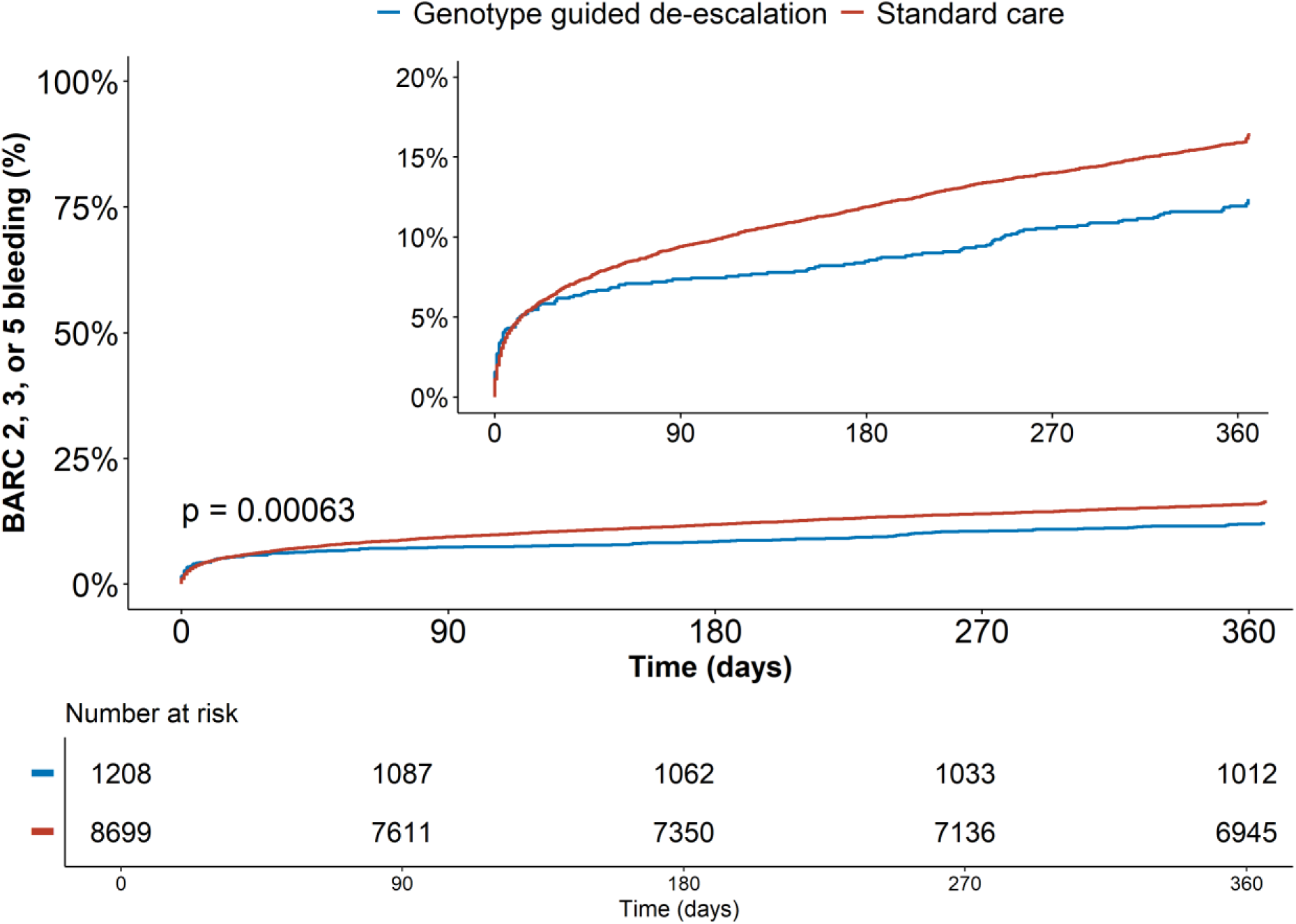
Kaplan-Meier curves for cumulative incidence of (A) the primary ischemic endpoint (cardiovascular mortality, myocardial infarction, or stroke), showing similar event rates between the genotype-guided cohort (blue) and standard care cohort (red), and (B) the primary bleeding endpoint (BARC 2, 3, or 5 bleeding), illustrating lower bleeding rates in the genotype-guided cohort (blue) with increasing divergence over time compared to standard care (red).

**Table 3.** Clinical outcomes of the genotype guided compared with the standard care cohort.

| Outcomes | Genotype<br>guided Cohort<br>(N = 1,207) | Standard Care<br>Cohort<br>(N = 8,699) | Unadjusted |  | Adjusted |  |
| --- | --- | --- | --- | --- | --- | --- |
|  |  |  | HR (95% CI) | P-Value | HR (95% CI) | P-Value |
| MACE | 107 (8.9%) | 897 (10.3%) | 0.85 (0.70-1.04) | 0.12 | 1.05 (0.85-1.29) | 0.64 |
| BARC 2, 3 or 5 | 146 (12.1%) | 1384 (15.9%) | 0.74 (0.63-0.88) | 0.0007 | 0.79 (0.67-0.94) | 0.01 |
| NACE | 137 (11.3%) | 1253 (14.4%) | 0.77 (0.655-0.92) | 0.004 | 0.91 (0.76-1.09) | 0.31 |
| All-cause death | 52 (4.3%) | 492 (5.7%) | 0.76 (0.57-1.01) | 0.05 | 1.08 (0.80-1.44) | 0.63 |
| CV death | 43 (3.6%) | 368 (4.2%) | 0.84 (0.61-1.15) | 0.27 | 1.30 (0.94-1.81) | 0.11 |
| MI | 44 (3.6%) | 423 (4.9%) | 0.74 (0.54-1.01) | 0.06 | 0.87 (0.63-1.19) | 0.38 |
| Stroke | 33 (2.7%) | 177 (2.0%) | 1.34 (0.92-1.94) | 0.12 | 1.46 (0.99-2.14) | 0.06 |
| Ischemic Stroke | 14 (1.2%) | 94 (1.1%) | 1.27 (0.72-2.24) | 0.41 | 1.56 (0.83-2.94) | 0.17 |
| Stent Thrombosis | 10 (0.8%) | 75 (0.9%) | 0.96 (0.48-1.85) | 0.90 | 0.81 (0.41-1.61) | 0.55 |
| BARC 2 | 124 (10.3%) | 1132 (13.0%) | 0.78 (0.64-0.93) | 0.007 | 0.83 (0.69-1.01) | 0.06 |
| BARC 3 or 5 | 31 (2.6%) | 326 (3.7%) | 0.68 (0.47-0.98) | 0.04 | 0.80 (0.55-1.17) | 0.25 |
| BARC 3 | 31 (2.6%) | 303 (3.5%) | 0.73 (0.50-1.06) | 0.09 | 0.85 (0.58-1.24) | 0.40 |

CV = cardiovascular; MI = myocardial infarction; BARC = Bleeding Academic Research Consortium; HR = hazard ratio; CI = confidence interval. \*Hazard ratios are based on an unadjusted model.

#### Secondary Outcomes

NACE occurred in 137 patients (11.3%) in the genotype guided cohort and 1,253 patients (14.4%) in the standard care cohort (_adj_HR, 0.91, 95% CI, 0.76-1.09; *P* = 0.31). The incidences of the individual components of all composite end point are shown in Table 2. The rate of death from any cause was 4.3% in the genotype guided cohort and 5.7% in the standard care cohort (_adj_HR, 1.08; 95% CI, 0.80-1.44, *P* = 0.63). The incidence of myocardial infarction was 3.6% in the genotype guided cohort and 4.9% in the standard care cohort (_adj_HR 0.87; 95% CI, 0.63-1.19, *P* = 0.38). There was no difference in the incidence of stent thrombosis (0.8% vs. 0.9%, _adj_HR 0.81, 95% CI, 0.41-1.61 *P* = 0.55).

Major bleeding (BARC 3 or 5) occurred in 2.6% of patients in the genotype guided cohort and 3.7% in the standard care cohort (_adj_HR, 0.80, 95% CI, 0.55-1.17; *P* = 0.25). The rate of BARC 2 bleeding was 10.3% in the genotype guided cohort and 13.0% in the standard care cohort (_adj_HR 0.83, 95% CI 0.69-1.01, *P* = 0.06).

### Sub-group analyses

Analyses of the primary outcomes were performed in specified subgroups. The results were generally consistent with those in the whole cohort (**Supplementary Figure 1**). In patients with high bleeding risk (HBR), based on a PRECISE-DAPT of 25 or higher, MACE occurred in 45 of 319 patients (14.1%) in the genotype guided cohort and 499 of 2,606 patients (19.1%) in the standard care cohort (_adj_HR, 0.85; 95% CI, 0.62-1.40; *P* = 0.31, *P*-value for interaction = 0.051). Major or non-major clinically relevant bleeding occurred in 52 of 319 patients (16.3%) in the genotype guided cohort and 572 of 2,606 patients (21.1%) in the standard care cohort (_adj_HR, 0.80; 95% CI, 0.60-1.07; *P* = 0.14).

In patients with complex PCI, MACE occurred in 13 of 172 patients (7.6%) in the genotype guided cohort and 125 of 1,010 patients (12.4%) in the standard care cohort (_adj_HR, 0.77; 95% CI, 0.43-1.40; *P* = 0.39), and major or non-major clinically relevant bleeding occurred in 25 of 172 patients (14.5%) in the genotype guided cohort and 178 of 1,010 patients (17.6%) the standard care cohort (_adj_HR, 0.99; 95% CI, 0.64-1.53; *P* = 0.97).

### Sensitivity analyses

After propensity score matching, 1,207 genotype-guided patients were matched to 3,441 standard care patients, with successful matching confirmed by standardized mean differences below 0.10 (**Supplementary Appendix Table 2)**. Results were consistent with those in the overall cohort (**Supplementary Appendix Table 3** and **Supplementary Figure 2**), showing a significant reduction in major or non-major clinically relevant bleeding (12.0% in the genotype guided cohort vs. 16.6% in the standard care cohort, _adj_HR, 0.73; 95% CI, 0.61-0.87, *P* = 0.0006) and comparable event rates for MACE (8.9% in the genotype guided cohort vs. 8.6% in the standard care cohort, _adj_HR, 1.04; 95% CI, 0.83-1.30, *P* = 0.74)

Sensitivity analyses excluding patients on oral anticoagulants (analysing 1,028 genotype-guided vs. 7,276 standard care), those not following the recommended P2Y12-inhibitor (analysing 988 genotype-guided vs 8,699 standard care) and only including patients undergoing PCI (analysing 905 genotype-guided vs. 5,991 standard care), all showed consistent results (**Supplementary Appendix Table 4**, **5** and **6**).

## Discussion

This study assessed the clinical impact of the implementation of a *CYP2C19* genotype-guided de-escalation strategy in routine practice. At 12 months, among patients admitted with ACS and receiving antiplatelet therapy, the genotype-guided approach significantly reduced major and non-major clinically relevant bleeding compared to standard care. There was no evidence of an increase in the rate of the combined ischemic endpoint, a composite of cardiovascular death, MI, or stroke, between the genotype-guided and standard care cohorts. In addition, the genotype-guided approach significantly reduced P2Y12-inhibitor treatment alterations and dyspnea-related discontinuations compared to standard care, indicating improved adherence and a lower burden of adverse effects. These findings build on our previously published analysis, which evaluated a smaller ACS population lacking power to detect clinical differences.[16] In contrast, the current analysis provides sufficient power to confirm the safety and effectiveness of genotype-guided therapy in routine practice.

The challenge of antiplatelet therapy in patients with ACS lies in achieving an optimal balance between effective platelet inhibition and the minimization of bleeding risk, without compromising protection against ischemic events. Common strategies aiming to achieve this, include shortening the duration of DAPT by discontinuing either aspirin or the P2Y12-inhibitor and continuing single antiplatelet therapy, or de-escalation, whether guided or unguided.[17] In genotype-guided de-escalation, genetic testing is used to identify a patient’s *CYP2C19* metabolizer status, allowing clinicians to safely switch normal metabolizers from a more potent P2Y12-inhibitor, such as ticagrelor or prasugrel, to clopidogrel. However, determining the optimal approach between a genotype-guided de-escalation strategy and P2Y12 monotherapy following a short period of DAPT is challenging, as no direct comparisons have been made in clinical trials. The current ESC ACS guidelines provide a Class IIb, Level of Evidence A recommendation for P2Y12 receptor inhibitor de-escalation as an alternative strategy to reduce bleeding risk.[2] However, specific recommendations whether de-escalation should be guided or unguided are lacking. Interestingly, the guidelines advise against de-escalation of antiplatelet therapy within the first 30 days after an ACS event but do not cite supporting studies for this recommendation. While very early discontinuation of DAPT (within thirty days after ACS or PCI) has been linked to increased ischemic outcomes, none of the early de-escalation strategies, whether guided or unguided, have demonstrated such risks to date.[9,18–20] Our findings align with those of the POPular Genetics trial, highlighting the safety of early de-escalation (preferably within <48 hours) upon admission, demonstrating no observed increase in ischemic events in the genotype-guided cohort. Additionally, Kaplan-Meier curve analysis of our trial reveals no elevated risk during the first 30 days of follow-up. While subgroup analyses consistently showed a reduction in bleeding across all subgroups, the borderline non-significant P-value for interaction suggests a potential modification of the treatment effect by HBR status. However, MACE event rates were similar in non-HBR patients (7.0% vs. 6.5%), with confidence intervals crossing one. Notably, the non-HBR subgroup included a large number of patients (888 in the genotype-guided cohort vs. 6,093 in the standard care cohort), underscoring the statistical power to detect meaningful differences. Based on these results, we conclude that a genotype-guided de-escalation strategy during initial hospital admission is safe for all patients.

While this study represents the largest real-world implementation of a *CYP2C19*-guided de-escalation strategy, other large-scale initiatives have successfully integrated *CYP2C19* genotype-guided antiplatelet therapy into clinical practice. These initiatives often employ escalation strategies, where clopidogrel is used by default, and patients with *CYP2C19* LOF alleles are escalated to ticagrelor or prasugrel. For example, a study by the IGNITE Network, which included an cohort of 3,342 patients across nine United States centres, reported an increased risk for adverse cardiovascular events— including death, MI, ischemic stroke, stent thrombosis, or hospitalization for unstable angina—with clopidogrel in patients carrying a LOF allele.[21] Importantly, no difference in cardiovascular risk was observed between clopidogrel and alternative therapies in patients without a LOF allele, aligning with the results from our study and that of the POPular Genetics.[22] This data from the IGNITE network demonstrate that escalation strategies can improve ischemic outcomes without increasing bleeding events, in line with the results of the TAILOR-PCI trial.[23] Although some regard TAILOR-PCI as a negative study, it provided compelling evidence that escalation strategies can optimize patient outcomes, particularly during the first months post-PCI.[23,24] Additionally, a pre-specified analysis of cumulative ischemic events further supports the benefits of this strategy.[25]

While escalation strategies may reduce ischemic events, their cost-effectiveness is limited, as the default choice, clopidogrel, is significantly more affordable than ticagrelor. In contrast, the cost-saving potential of a de-escalation strategy is more substantial, as instead of treating every patient with ticagrelor/prasugrel as the default, around 70% can be de-escalated to the cheaper clopidogrel.[8,26] Nonetheless, the broader implementation of this strategy still faces notable challenges. Currently, hospitals bear the cost of genetic testing, while insurers benefit from reduced medication expenses. This imbalance limits adoption, as hospitals are unlikely to fund testing independently. National reimbursement of genetic testing could address this issue, enabling broader implementation and amplifying the cost-saving potential through economies of scale. Further integration requires addressing logistical barriers such as accessibility and turnaround time. Collaborative efforts between researchers, clinicians, and policymakers are critical to ensure equitable access.

A key challenge in optimizing antiplatelet therapy is that variability in response to clopidogrel extends beyond genetic factors. The ABCD-GENE score, which incorporates age, BMI, diabetes, kidney function, and *CYP2C19* genotype, provides a comprehensive tool for stratifying patients at risk for high platelet reactivity (HPR) and adverse ischemic outcomes.[27] Patients with an ABCD-GENE score ≥10 showed trends toward improved outcomes with alternative P2Y12-inhibitors compared to clopidogrel, particularly among carriers of *CYP2C19* LOF alleles.[28] This highlights the necessity for tailored approaches, balancing patient-specific ischemic and bleeding risks. Integrating genetic results within the broader clinical context, including procedural complexity and patient comorbidities, will enhance the precision of antiplatelet therapy strategies.

A key strength of our study is the demonstration of comparable outcomes for both ischemic and bleeding events to those observed in the POPular Genetics trial. The hazard ratios for BARC 2, 3, or 5 bleeding closely align with our adjusted and unadjusted models. Notably, the sensitivity analysis in PCI patients only confirmed consistent bleeding reduction, reinforcing the efficacy of a genotype-guided strategy in ACS patients undergoing PCI. Furthermore, our findings underscore the importance of implementation studies, as randomized trial populations are typically younger and at lower risk. Our study enrolled an older population (66 vs. 62 years) with higher mortality (5.5% vs. 1.5%), better reflecting real-world conditions. Our results further demonstrate that de-escalation is safe and effective in a broad ACS population, unlike the POPular Genetics trial, which focused solely on STEMI patients. An important observation is that approximately 29% of patients in the standard care group received clopidogrel, nearly half of whom were also treated with oral anticoagulation, compared with only 7% in the POPular Genetics trial. This high clopidogrel use would theoretically attenuate bleeding differences between groups, yet we still observed a bleeding reduction comparable to that seen in POPular Genetics. This finding likely reflects the inherent limitations of the observational study design but also supports the robustness of the bleeding benefit associated with genotype-guided de-escalation.

### Limitations

We recognize several limitations of our study. Our study’s real-world design allowed physicians to decide whether to follow genotype recommendations, introducing variability in adherence to the strategy. Also, due to the non-randomized nature of the study, this resulted in baseline differences between both cohorts that could potentially confound the results. While adjustment methods, including propensity score matching, successfully balanced both cohorts, residual confounding related to the selection of antiplatelet therapy cannot be entirely ruled out. The observed differences in hazard ratios before and after adjustment indicate that part of the observed association was influenced by confounding. However, after adjusting for confounders, our primary findings remained consistent. While the strategy is implemented in multiple hospitals, the majority of most patients in the genotype guided group were enrolled in one site. Patients in the other sites were more often later de-escalated due to offsite genotyping. While most patients still were de-escalated within three weeks, it may have affected ischemic and bleeding outcomes during this period. Nevertheless, it is unlikely that this has affected the overall results of the study since offsite genotyping was only used in a minority of the patients. Lastly, a considerable proportion of patients were treated with oral anticoagulation, reflecting the broader ACS population. Although triple therapy use was higher in the standard care cohort, potentially influencing bleeding risk, we adjusted for this in our models. Moreover, sensitivity analyses excluding patients on oral anticoagulation confirmed consistent results.

## Conclusion

In conclusion, in patients with ACS receiving antiplatelet therapy, implementing implementation of a *CYP2C19* genotype-guided de-escalation strategy in clinical practice significantly reduced major and non-major clinically relevant bleeding compared to standard care at 12 months, without an increase in ischemic events, including cardiovascular death, MI, or stroke.

## Acknowledgements

None.

## Data availability statement

The data underlying this article will be shared on reasonable request to the corresponding author.

